# First trimester maternal serum microRNA expression profile differentiates between uncomplicated pregnancies, and pregnancies which develop pre-eclampsia

**DOI:** 10.1101/2023.05.09.23289708

**Authors:** Paula L. Hedley, Severin Olesen Larsen, Karen R. Wøjdemann, Anne-Cathrine Shalmi, Sophie Placing, Line Rode, Anne Catrine Gjerris, Karin Sundberg, Ann Tabor, Michael Christiansen

## Abstract

Numerous circulating microRNAs (miRNAs) have been detected in maternal blood. Initial studies in third trimester demonstrated differential miRNA expression profiles between uncomplicated pregnancies and pregnancies complicated by pre-eclampsia (PE). Recently, studies in first trimester have shown similar differential profiles, however, these studies were often under-powered. We conducted a nested case-control study, in which serum samples, taken between 10-14 weeks gestation, were obtained from 413 singleton pregnant women, 126 of which later developed PE. Total RNAs were purified and a selection of 46 miRNAs plus two miRNA controls were quantitated by real time quantitative PCR. Seven of the miRNAs, hsa-miR-181b-5p, -323a-3p, -518b, -363-3p, -20a-5p, -29a-3p, and -142-3p, could differentiate between uncomplicated pregnancies and pregnancies which develop PE, but only a single miRNA, hsa-miR-363-3p, could differentiate between mild and severe PE. A combination of all seven differentiating miRNAs was the best at discriminating between PE and uncomplicated pregnancies (AUC= 0.879). In conclusion, first trimester maternal serum miRNA expression profile could differentiate between uncomplicated pregnancies and pregnancies complicated by PE. These circulating miRNA markers have the potential to improve risk assessment of PE in the first trimester, weeks before the onset of symptoms.

## Introduction

Pre-eclampsia (PE) is a major cause of maternal and foetal morbidity and mortality worldwide. Characterized by hypertension, frequently associated with proteinuria [1, 2], PE can damage multiple organ systems and affects approximately 2-8% of pregnancies globally [3, 4]. The onset of PE symptoms occurs most often in the third trimester, but can occur any time after 20 weeks of gestation. A considerable proportion of PE pregnancies require preterm delivery, with the risks associated with prematurity for the child [5, 6]. PE is associated with cardiovascular and other morbidity postnatally for both mother and child [7, 8]. Treatment with aspirin may reduce the risk of preterm PE if initiated prior to gestational week 16 [6, 9]. PE is subclassified into early- or late PE and whether it is associated with fetal growth restriction or not, and this heterogeneity may reflect different etiologies [10]. Early inadequate uteroplacental perfusion is considered the hallmark of PE [11], leading -ultimately - to maternal and fetal syndromes through endothelial dysfunction [12]. However, despite decades of study, the identification of maternal risk factors [13] and biomarkers [13, 14], and the design of first trimester risk algorithms that reasonably identify preterm pregnancies [12, 15], there is still a need for improving early pregnancy prediction models for PE and its various clinical presentations.

Since their discovery in the late 1990’s microRNAs (miRNAs), a class of small (∼22nts) non-coding RNAs have been reported as a potentially important class of biomarkers for several diseases [16]. MiRNAs serve a post-transcriptional regulatory function, typically, translational repression or degradation, with respect to their target mRNAs [16]. Cell and tissue specific miRNA expression profiles as well as their ability to be transported within and between cells enable miRNAs to be involved in diverse physiological and pathophysiological processes [17, 18]. At the time of writing, there are more than 1,900 human miRNAs published in miRBase, a searchable database of published miRNA sequences and annotation [19].

Circulating miRNAs (also known as extracellular miRNAs), have been extensively studied in body fluids, such as plasma, serum, urine, saliva, cerebrospinal fluid, amniotic and follicular fluids [20]. These circulating miRNAs may be passively released from damaged cells or dying cells or they may be secreted bound to proteins [21, 22], or within extracellular vesicles [23]. Altered circulating miRNA expression profiles have been detected in patients with various diseases or disease stages [24]. Circulating miRNAs constitute useful biomarkers of disease and disease course and as they can be measured through non-invasive procedures may supplement or be preferable to current standard invasive tests, e.g. biopsies and chorion villous sampling [24].

Appropriate trophoblast proliferation is essential for normal placentation and pregnancy health in first trimester and human villous trophoblasts secrete placenta-specific miRNA into the maternal circulation via exosomes [25]. Consequently, numerous circulating miRNAs have been detected in maternal blood, and it has been demonstrated that their expression profile varies throughout pregnancy and between uncomplicated pregnancies and pregnancies with adverse outcomes [26]. Several studies have reported altered miRNA expression profiles in plasma and serum from women whose pregnancies were complicated with PE [27-34]. First trimester circulating miRNA markers have the potential to identify at risk pregnancies weeks before the onset of symptoms [29, 34-40].

Previous studies of circulating miRNAs in PE were mostly conducted with small sample sizes, and/or the samples were typically collected after onset of PE symptoms or following delivery [41]. Our study was performed on a comparatively large number of cases and controls (413 singleton pregnant women, of which 126 later developed PE) and serum samples were collected during the first trimester (10-14 weeks). A panel of 46 miRNAs which are mostly pregnancy or pregnancy disorder related were investigated in this study [26, 35, 42-51]. Our hypothesis is that the first trimester serum miRNA expression profile will be able to differentiate between uncomplicated pregnancies, and pregnancies complicated by PE. This differential profile may improve prognosis and diagnosis of PE in early stage of pregnancy.

## Results

The profile of serum miRNA was investigated with a panel of 46 miRNAs (Supplementary Table 1) and 413 first trimester serum samples from either uncomplicated pregnancies (n = 287), or pregnancies (n = 126) which later developed either mild PE (n = 98) or severe PE/HELLP syndrome (n = 28). Four miRNAs (hsa-miR-181b-5p (P <0.0001), hsa-miR-323a-3p (P <0.0001), hsa-miR-518b (P <0.0001), and hsa-miR-363-3p (P <0.0001)) were up-regulated and three miRNAs (hsa-miR-20a (P <0.0001), hsa-miR-29a (P <0.0001), and hsa-miR-142-3p (P = 0.0004)) were down-regulated in pregnancies which developed PE (mild or severe) compared to uncomplicated pregnancy (Figure 1). Only hsa-miR-363-3p produced a statistically significant different expression level between mild and severe PE (P = 0.005) (Figure 1).

**Figure 1:**
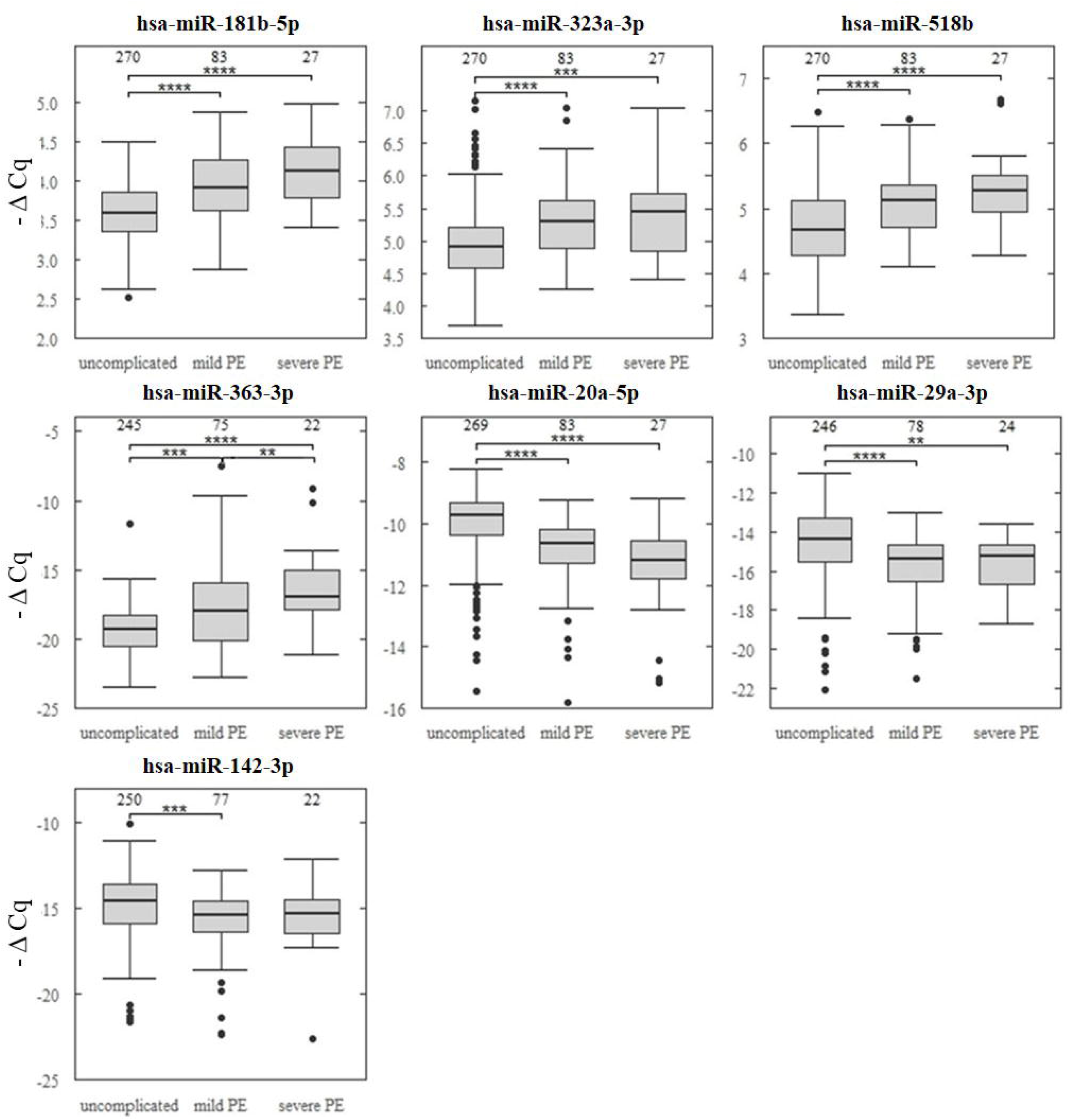
Boxplots of delta Cq values for the seven miRNAs determined to have significantly different expression levels between uncomplicated pregnancies and pregnancies which went on to develop PE (mild or severe). Number of samples assessed per group is listed above each box. *P<0.05, **P<0.01, ***P<0.001, ****P<0.0001.

Logistic regression analysis determined maternal weight to be significantly associated with developing PE (OR 1.04, 95 % CI 1.02, 1.06) (Table 1). Receiver operator curves (ROC), adjusted for maternal weight, of the individual miRNA biomarkers found that hsa-miRNA-20a-5b (area under the curve (AUC) = 0.779), 181b-5p (AUC = 0.753) and 363-3p (AUC = 0.728) discriminated PE from uncomplicated pregnancy markedly better than the other miRNA biomarkers (Figure 2). However, a combination of all seven miRNA biomarkers, adjusted for maternal weight, discriminated PE from uncomplicated pregnancies best (AUC = 0.879) (Figure 2).

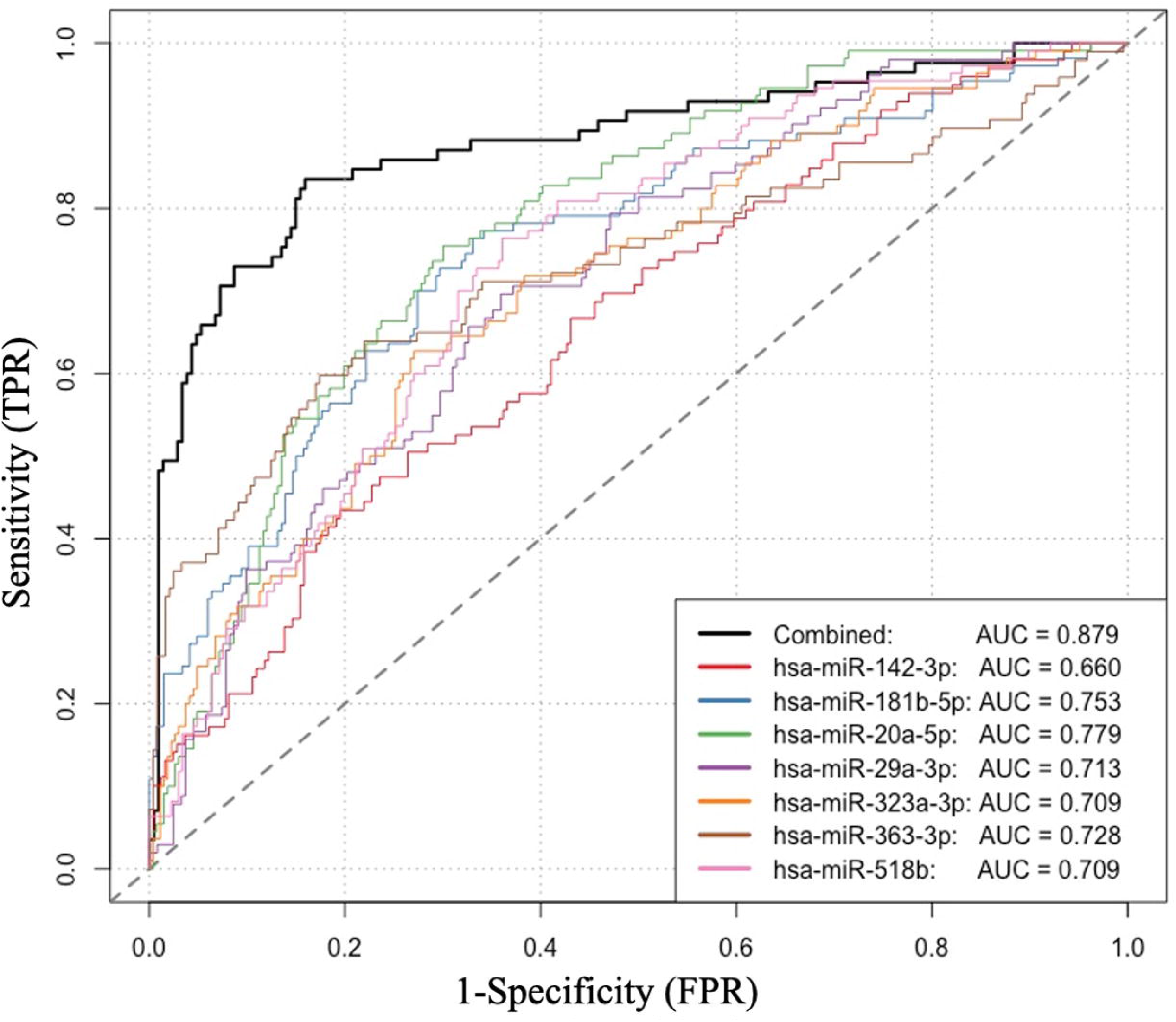

|  | **Controls (N = 287)** | **PE (N = 126)** | **OR (95% CI)** |
| --- | --- | --- | --- |
|  | *median (IQR)* | *median (IQR)* |  |
| GA test (days) | 90 (88 - 94) | 90.5 (87 - 93) | 0.97 (0.89 - 1.07) |
| Maternal age (years) | 30 (27 - 33) | 30 (27 - 33) | 1.01 (0.96 - 1.06) |
| Maternal weight (Kg) | 62 (57 - 70) | 68 (62 - 69.5) | 1.04 (1.02 - 1.06) *** |
| Crown-Rump length (mm) | 67 (61 - 74) | 67 (61 - 72) | 1.00 (0.96 - 1.06) |
|  | *mean (sd)* | *mean (sd)* |  |
| Parity | 1.70 (0.97) | 1.70 (1.11) | 0.94 (0.76 - 1.17) |

## Discussion

Pregnancy involves many complex physiological events in which miRNAs might play a role. Embryo implantation and placentation are two critical steps for establishing pregnancy [52], however, current knowledge regarding the physiology of human placental cell proliferation, invasiveness, differentiation and apoptosis during the first trimester is limited. Research into the regulation of trophoblast activities by miRNA may help to address the pathophysiological basis of pregnancy disorders.

In this study expression profiles of 46 first trimester maternal serum miRNAs, mostly pregnancy or pregnancy disorder related, were investigated by RT-qPCR among 270 uncomplicated pregnancy, and 110 pregnancies which would go on to develop PE. We found seven miRNAs (hsa-miR-181b-5p, -323a-3p, -518b, -363-3p, -20a-5p, -29a-3p, and -142-3p) able to discriminate between uncomplicated pregnancies and pregnancies which would go on to be complicated by PE, but only hsa-miR-363-3p was able to discriminate between mild PE and severe PE. Several of these dysregulated miRNAs are known to influence placental functions e.g. trophoblast migration (hsa-miR-181b-5p and -20a-5p) [53, 54], invasion (hsa-miR-181b-5p) [54], differentiation (hsa-miR-20a-3p) [53], and proliferation (hsa-miR-518b, -20a-5p, and -142-3p) [46, 53, 55], as well as angiogenesis (hsa-miR-20a-5p) [56] and apoptosis (hsa-miR-29a-3p) [53]. Furthermore, hsa-miR-518b, -20a-5p, and -29a-3p have been found to be upregulated, and hsa-miR-363-3p has been found to be downregulated, in either serum and / or placentae from women with PE [57], while hsa-miR-323-3p has been found to be a biomarker of ectopic pregnancy [51].

Several methods, such as next generation sequencing, nanomaterial-based methods, microarrays, qRT-PCR, and other nucleic acid amplification-based methods have been developed to detect miRNA expression [58, 59]. Despite the promise of these methods, qRT-PCR remains the gold standard in miRNA detection techniques [59]. However, the use of different methodologies in the field produces results which are difficult to compare. Combined with often very small sample sizes and a lack of standardisation regarding the collection, preparation and extraction of samples and analytical methods for miRNA analysis [60], e.g. there are no universally accepted internal controls for miRNA analysis, makes miRNA studies especially challenging. It is, therefore, not surprising that we see contradictory findings in studies of miRNA expression in PE. Furthermore, studies of miRNAs in PE frequently collect samples late in pregnancy, after the onset of symptoms, or during delivery [61, 62]. We found that hsa-miR-518b was upregulated in first trimester maternal circulation, thus confirming Hromadnikova et al’s first trimester study findings [35]. However, our finding that hsa-miR-20a-5p and -29a-3p were both downregulated contradicts the findings of Jairajpuri et al [63] and Wang et al [56]. Jairajpuri et al, found that hsa-miR-29a-3p was upregulated in plasma samples taken from severe PE patients, these patients had developed PE and it is possible that the miRNA profile changes between first trimester and the development of symptoms. Additionally, this study compared eight severe PE patients with 11 normotensive women which may impact the power of this study [63]. Wang et al, found that hsa-miR-20a-5p was upregulated in placental tissue of severe PE patients [56], again these samples were taken at delivery and the miRNA profile may not be comparable to the first trimester miRNA profile in maternal circulation. Furthermore, this study too may be underpowered with 10 severe PE patients being compared to 10 normotensive women [56].

Large-scale prospective longitudinal studies in well-defined populations are necessary to further investigate the clinical utilities of various circulating miRNAs during early stage of pregnancy. The dysregulation of miRNA’s, measured in maternal circulation during the first trimester, indicates that placental dysfunction leading to PE can be ascertained before the onset of symptoms, and may be useful as a risk assessment tool. A combination of the miRNAs identified in our study and other first trimester ultrasound and serum protein biomarkers [64-67], metabolic and physiological parameters [68-73] may greatly improve early risk assessment of PE, and, in consequence, improve prophylaxis of PE, decreasing the health burden of the disease and its associated morbidities.

## Material and Methods

### Study design

This study was designed as a nested case-control sub-study, nested within the Copenhagen First Trimester Screening Study Cohort, collected from 1997 to 2001 [74].A number of studies on Down syndrome markers and PE markers have been conducted using this sample set [68, 71, 72, 75, 76]. All samples were drawn during the first trimester (weeks 10-14).

### Serum samples

Serum samples were selected from 126 cases (aged 18-46 years) who went on to develop PE with proteinuria (98 mild PE and 28 severe PE/HELLP syndrome, classified according to clinical and paraclinical criteria from the Danish Society of Obstetrics and Gynecology [77]), and 287 (aged 19-43 years), parity, and gestational age at sampling matched controls who went on to have uncomplicated pregnancies. Basic clinical information is given in Table 1. All samples were collected in dry containers and kept at 4 ⍰C for a maximum of 48 h until shipment to the Statens Serum Institut. Samples were subsequently stored at −20 ⍰C. The samples had been subjected to several freeze-thaw cycles prior to the measurement of miRNAs.

### RNA purification to real time qPCR

The expression profile of 48 miRNAs (supplementary Table 1) were generated for all 423 serum samples. A variable number of analyses failed or insufficient material was present, consequently, the number of results per miRNA species is given in Figure 1.

The RNA purification and real time quantitative PCR (RT qPCR) were performed as previously described [78]. Briefly, total RNA (including microRNAs) was purified by Norgen total RNA purification kit (Norgen Biotek Corp., Ontario, Canada) from 100 μl serum. 10 mM dithiothreitol and 1.7 pM of *C. elegans* synthetic miRs-39, -54, and -238 (Tag Copenhagen A/S, Denmark) were added to a volume of lysis buffer sufficient for all the samples. This volume was then aliquoted out into 2 ml portions and kept at −20 °C until used. The RNA was purified according to the instructions of the Manufacturer. Before elution of RNA 1 μl of RNAse inhibitor (Applied Biosystems, Foster City, CA, USA, 20 U/μl) was added to every elution tube.

Reverse transcription (RT) was performed by the TaqMan microRNA RT kit (Applied Biosystems, Foster City, CA, USA) with modification: RT reaction volume was 10 μl using 1 μl of MultiScribe™ reverse transcriptase, 3 μl of RT primer-mix (The RT-primer-mix consisted of equal volumes of each of 48 different 5X RT microRNA-specific stem-loop primers, Supplementary Table 1), 1 μl of 10X buffer, 0.2 μl of 100 mM dNTPs, 0.15 μl of RNAse inhibitor and 4.65 μl of the purified RNA. RT was performed on an ABI 2720 Thermal Cycler (Applied Biosystems, Foster City, CA, USA) at 16 °C for 30 min, 42 °C for 30 min, 85 °C for 5 min, then hold at 4 °C.

Specific target amplification was accomplished using the TaqMan PreAmp master mix and a mix of 48 TaqMan MicroRNA Assays (Applied Biosystems, Foster City, CA, USA). The pre-amplification mixture (10 μL) contained 2.5 μL of diluted cDNA (diluted 1:3 with H2O), mixed with 5 μL of 2X TaqMan PreAmp master mix and 2.5 μL of the 0.2X TaqMan miR-assay mix. Pre-amplification was performed at 95 °C for 10 min, 16 cycles of (95 °C, 15 s; 60 °C, 4 min), then hold at 4 °C.

Pre-amplified samples (diluted 1:5) and 20X TaqMan miR assays were applied to primed 96.96 dynamic array chips using loading and assay reagents according to the instructions of manufacturer (Fluidigm Corp., CA, USA). All 48 miRNA-assays were performed in duplicate. RT qPCR was performed in a BioMark system (Fluidigm Corp., CA, USA) using single probe (FAMMGB, reference: ROX) settings and the GE 96×96 standard v.1 protocol with 40 cycles. Data were processed with the Fluidigm real-time PCR analysis software (v. 3.1.3) using the auto-detectors setting.

### Data handling

Average raw Cq scores above 35 were considered as out of the limit of detection. Average Cq values were first subtracted from the average Cq of the two Cel-miRs (cel-miR-54 & 238) for that sample to correct technical variance. The yielded -ΔCq (average Cq of the two Cel-miRs - average Cq of miR for that sample) values were used in further analyses. All -ΔCq values then were further normalized with the average -ΔCq of 10 microRNAs (hsa-miR-1, 20a-5p, 126-3p, 155-5p, 181b-5p, 222-3p, 223-3p, 299-5p, 323a-3p, 518b) detected in all samples to correct variations in total input RNA. These further normalized expression values were used for the statistical analyses.

### Statistical analysis

Statistical analyses were performed using R software (version 4.0.3). Statistical significance was determined by Kruskal-Wallis with Dunn’s post hoc test and Benjamini-Hochberg correction. Gestational age at time of testing, maternal age, maternal weight, parity, and crown-rump length were assessed by logistic regression to determine which foetal/maternal factors were associated with developing PE. The ROCs, adjusted for maternal weight, was used to evaluate the diagnostic sensitivity of first trimester serum miRNAs in pregnancies which went on to develop PE. The AUCs were calculated and compared. The p-value < 0.05 represents statistically significant change.

## Data Availability

Data are not publicly available to preserve the privacy of participants under the European General Data Protection Regulation.

## Supplementary Materials

**Supplementary Table 1:**
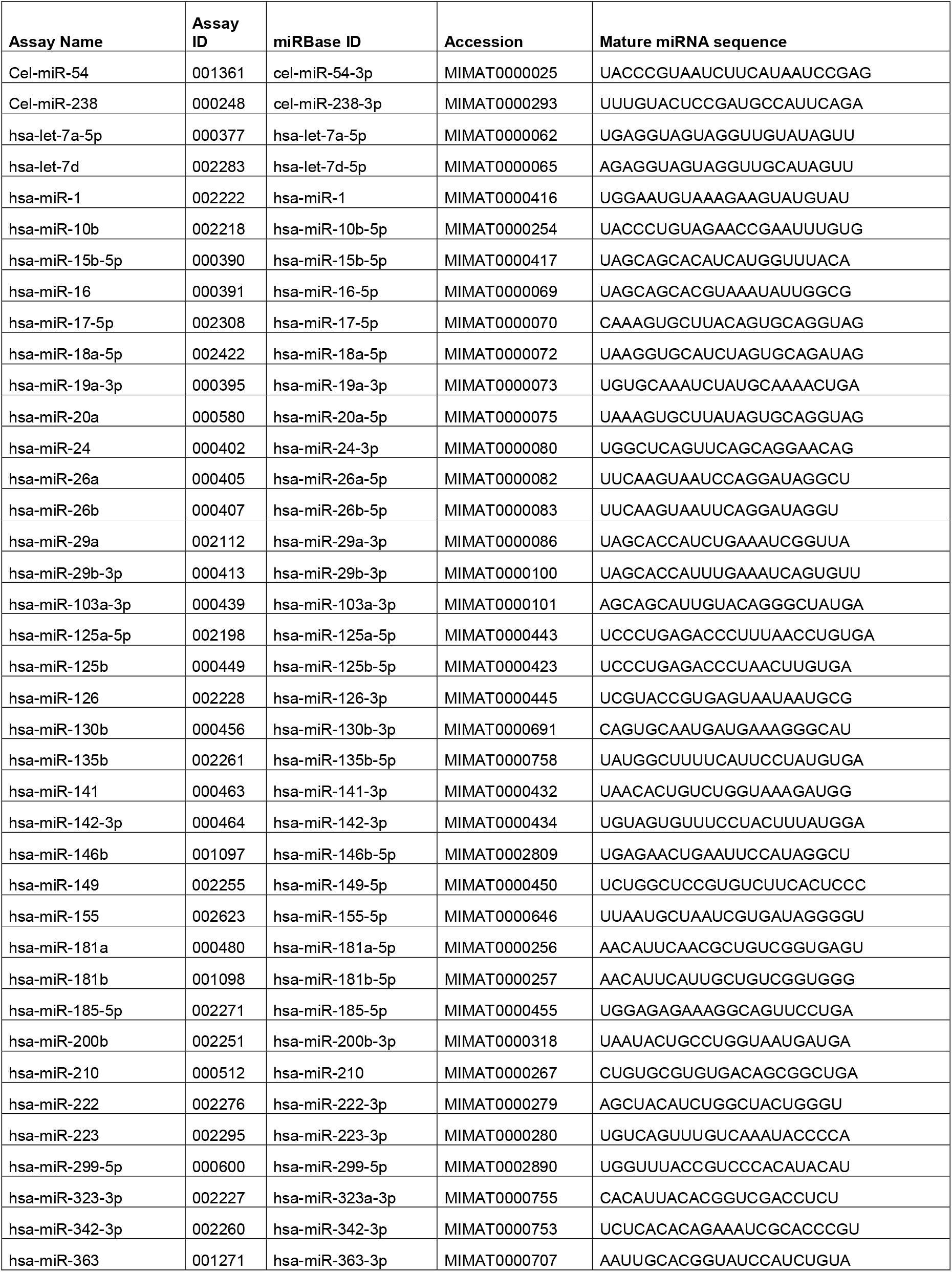
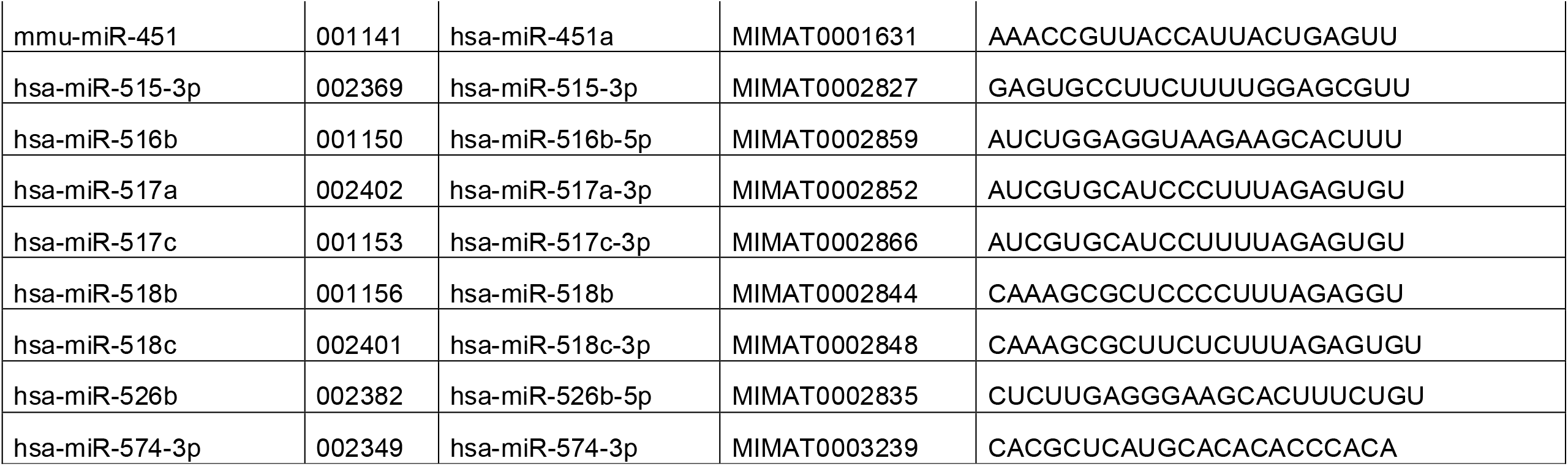
48 miRNA Assays. Cel-miR-54 & - 238 are technical controls.

## Acknowledgements

We would like to acknowledge the efforts of Dr Anting Liu Carlsen who contributed to the design of the study, performed the experimental work, and wrote the first draft of this manuscript. Unfortunately, Dr Liu Carlsen passed away before the manuscript could be completed. Dr Carlsen’s contribution to this work has been listed in the author contributions statement under A.L.C.

This research has been conducted using the Danish National Biobank resource, supported by the Novo Nordisk Foundation.

## Author Contributions

Conceptualization, A.L.C., P.L.H. and M.C.; Methodology, A.L.C., S.O., P.L.H. and M.C; Formal Analysis, A.L.C., P.L.H., M.C., S.O.; Resources, K.R.W., A.C.S., S.P., A.C.G., K.S., A.T. and M.C.; Data Curation, A.L.C., P.L.H. and M.C.; Writing – Original Draft Preparation, A.L.C. and P.L.H.; Writing – Review & Editing, All authors

## Ethics Approval Statement

The Copenhagen First Trimester Screening study was conducted according to the guidelines of the Declaration of Helsinki and approved by the Scientific ethics committee of the cities of Copenhagen and Frederiksberg (No. (KF) 01-288/97) and the Data Protection Agency.

